# Predictors of positive remodeling in patients with acute ischemia stroke

**DOI:** 10.1101/2024.03.30.24305124

**Authors:** Gen Li, Di-hao Xu, Jing-wen Xie, Li-ping Ma, Min Shu, Zhi-bin Zeng

**Affiliations:** Emergency Department, Huazhong University of Science and Technology Union Shenzhen Hospital, Shenzhen, Guangdong, China; Radiology Department, Huazhong University of Science and Technology Union Shenzhen Hospital, Shenzhen, Guangdong, China

**Keywords:** vascular remodeling, intracranial atherosclerosis, acute ischemia stroke, high-resolution vessel wall imaging

## Abstract

**Objective:** This study aimed to delineate the differences in plaques characteristics between positive remodeling (PR) and non-positive remodeling (NPR) in patients with Acute ischemia stroke (AIS), and to assess the associated relative risk factors for PR in intracranial atherosclerosis (ICAS).

**Methods:** Patients with AIS were enrolled and underwent high-resolution magnetic resonance vessel wall imaging (HR-VWI) within two weeks of symptom onset. We compared plaque morphologies and clinical variables between the PR and NPR cohorts. Binary logistic regression was employed to isolate independent predictors of PR, and the predictive model’s sensitivity and specificity were gauged using the receiver operating characteristic curve.

**Results:** A total of 84 patients (mean age 58.07±1.35 years, 78.6% male) were categorized into PR (n=28, 33.3%) or NPR (n=56, 66.4%) groups based on the remodeling index (RI). The PR group exhibited a significantly higher percent plaque burden (P<0.001), wall area (WA) (P<0.001), plaque length (P=0.018), and RI (P<0.001), alongside elevated fasting blood glucose (P=0.01) and a higher prevalence of hypertension and diabetes mellitus (P=0.16, 0.025). Logistic regression analysis identified percent plaque burden as an independent predictor of PR (odds ratio, 4.19, per 10% increase; [95% CI, 1.79-9.8]; *P*=0.001).

**Conclusion:** HR-VWI is an effective tool for identifying PR in AIS caused by ICAS, with a high plaque burden being an independent correlate of PR.

## Introduction

Intracranial atherosclerosis (ICAS) is the primary cause of acute ischemic stroke (AIS), characterized by focal vascular inflammation and lipid accumulation within plaques^1,2^. Arterial remodeling is a compensatory response to progressive plaque accumulation in the vessel wall, often manifesting as changes in the outer wall size to maintain homeostasis and adapt to physiological demands^3-6^.

Vascular remodeling in atherosclerosis is a dynamic process that typically involves endothelial dysfunction, proliferation and migration of vascular smooth muscle cells (VSMCs), their phenotypic transformation, apoptosis, and extracellular matrix degradation^7,8^. Recent evidences also indicate a complex regulatory role for the adventitia in this process^9^. Positive remodeling (PR), or compensatory outward bulging of the vessel wall, may result from two possible biological mechanisms^10-14^: (1) Endothelium-dependent dilation mediated by local shear stress and (2) external bulging of the plaque following degradation of the underlying media and adventitia, or out migration of VSMCs and ischemic medial atrophy. Conversely, the “negative remodeling”, is the restriction of the vessel size or vascular wall degeneration, which reflects contraction of VSMCs or vessel fibrosis^8,15,16^. The significance of arterial remodeling in compensating for or exacerbating luminal stenosis in atherosclerosis has been predominantly recognized through in vivo ultrasound imaging, postmortem analyses, or animal model studies^2,17-20^.

The investigation into ICAS remodeling has been limited due to a lack of detailed histopathological analyses and non-invasive imaging techniques. Conventional angiography, including MRA, CTA, and DSA, is restricted to assessing luminal stenosis. However, high-resolution vessel wall imaging (HR-VWI) provides in vivo, sub-millimeter assessment of the arterial wall, utilizing advanced blood and cerebrospinal fluid suppression techniques^21^. The use of 3D isotropic imaging technology with an isotropic voxel size between 0.5 to 0.7mm enables whole-brain coverage and multi-plane and curved-planar reformations^22^, minimizing artifacts from the tortuous intracranial vessels.

HR-VWI has gained prominence for its ability to characterize high-risk plaques and determine stroke etiology^23-25^, however, the role of remodeling in ICAS and its clinical implications remain poorly defined. The varied remodeling patterns evident in the pathological progression of de novo atherosclerosis raise questions about the determinants of remodeling’s extent and direction. We posit that both local and regional vascular characteristics, as well as individual factors, contribute to positive vascular remodeling. The culprit plaque, influenced predominantly by clinical factors, offers valuable insights into the impact of individual-related factors on remodeling. HR-VWI facilitates the quantification of local plaque changes, thus, this study aims to evaluate the potential risk factors for PR in ICAS.

## Materials and methods

This retrospective study was approved by the local institutional review board. Patients presenting with AIS within 7 days of symptom onset and exhibiting ICAS plaque at the proximal segment of the middle cerebral artery (MCA) or basilar artery (BA) were prospectively enrolled from January 2021 to April 2023. Inclusion criteria: (1) Age >30; for those aged 30-49, the presence of atherosclerotic plaques in major arteries or at least two risk factors such as hypertension, diabetes, hyperlipidemia, and smoking. (2) Admission within seven days of symptom onset, with diffusion-weighted imaging (DWI) confirming infarction in territories supplied by the stenotic artery. (3) Completion of 3D-VWI scan within 14 days of hospital admission. (4) High-quality images and comprehensive clinical and laboratory data were available for analysis. Written informed consent was obtained from all participants. Exclusion criteria encompassed: (1) Other stroke etiologies like arterial dissection, aneurysm, moyamoya disease, subclavian steal syndrome, or cardio-embolism. (2) Chronic infarcts or transient ischemic attacks (TIAs) not evident on DWI. (3) Significant extracranial artery stenosis exceeding 50%. (4) MRI contraindications including metal implants, claustrophobia, or pregnancy. (5) Renal impairment or known gadolinium allergy.

Clinical indicators such as age, gender, symptom onset, hypertension, diabetes, hyperlipidemia, smoking history, and demographic data, along with cardiovascular risk factors and laboratory values (including fasting blood glucose (FBG), cholesterol (CHOL), triglyceride (TG), low density lipoprotein (LDL) and high-density lipoprotein (HDL)), were recorded.

### MRI parameters

All qualifying AIS patients underwent imaging on a 3.0-T MRI (SIGNA Pioneer, GE Healthcare) with an 8-channel phased-array head coil to obtain a three-dimensional gradient-echo sequence for T1-weighted, T2-weighted, and contrast-enhanced T1-weighted images, in addition to magnetic resonance angiography (MRA). Protocol details are provided in the Table 1.

**Table 1.** Parameters of HR-VWI MRI sequences.

| MR scan parameters | 3D T1WI CUBE Pre<br>&Postcontrast | 3D T2WI CUBE | MRA |
| --- | --- | --- | --- |
| TR/TE, ms | 650/12 | 2502/118 | 3.4/18 |
| voxel size, mm<sup>3</sup> | 0.8×0.8×0.8 | 0.8×0.8×0.8 | 0.7×0.9×1.2 |
| Slice thickness (mm) | 0.8 | 0.8 | 1.2 |
| Echo train length | 36 | 96 | 1 |
| Flip angle (degrees) | Variable Flip angle flag | Variable Flip angle flag | 20 |
| Field of view(mm<sup>3</sup>) | 192×192 | 192×192 | 220×206 |
| Matrix(frequency×phase) | 240×240 | 240×240 | 320×256 |
| Pixel Bandwidth | 244 | 244 | 162 |
| Intensity correction | SCENIC | SCENIC | PURE |
| Acquisition time(s) | 369 | 274 | 194 |

TR, repetition time; TE echo time.

### Images processing and analysis

Image quality was assessed on a scale from 1 to 4, based on the signal-to-noise ratio of all HR-VWI images, with 1 being the worst and 4 being the best. A grade of ≥3, indicating clear visualization of most luminal and outer wall boundaries, met the study’s requirements.

Images processing was conducted at the Syngo.via workstation (SIEMENS Healthcare) using MR TWIST toolkit. Initial steps involved the automatic extraction of the centerline, determined by manually-set start and end points of the culprit plaque, including the proximal and distal reference sits. Subsequent multi-planar and curved-planar reformations were executed along this centerline. All MR data were analyzed and measured on cross-sectional and curved-planar images by two radiologists (D.H.X and L.P.M with 4 and 6 years of diagnostic experiences, respectively). Discrepancies were resolved by a senior radiologist (Z.B.Z. with 28 years of diagnostic experience in cardiovascular MR imaging).

## Definition of plaque characteristics and measurements

The culprit plaque was either the sole plaque in the artery supplying the acute infarct lesion or the most unstable plaque, as indicated by wall thickening or enhancement when multiple plaques (≥2) were present in the same artery. Measurements of the morphological data of the culprit plaque were taken at the most narrowed lumen (MNL) site. The reference site was defined as the nearest cross-section exhibiting the least plaque presence, situated either proximal or distal to the plaque. When the culprit plaque was elongated, proximal and distal site data were recorded and averaged for reference. Post-contrast images facilitated the delineation of measurements for lumen and outer vessel contours, as well as plaque length.

Severity of stenosis was calculated as: (1−lumen area of MNL/reference lumen area) ×100%. The wall area (WA) was computed as the vessel area minus the lumen area (ie, WA=vessel area− lumen area). Normalized wall index (NWI) was defined as: WA/ (WA+ lumen area) ×100%. Consider that different vessel size of BA and MCA can be a potentially confounding parameter, plaque burden is therefore estimated by percent plaque burden, calculated as (WA_MNL_ /WA_reference_) ×100%. The ratio has been mentioned in previous studies^2,26,27^. The remodeling index (RI) is the ratio of the vessel area at the MNL site to the vessel area at the reference site. Positive remodeling is defined as an RI > 1.05, while negative remodeling (NPR) is indicated by an RI < 1.05. Intra-plaque hemorrhage (IPH) is identified by a signal intensity exceeding 150% of that of adjacent gray matter on T1-weighted images (T1WI). The extent and pattern of plaque enhancement were primarily assessed in relation to the normal-appearing intracranial arterial wall and the pituitary gland. The plaque enhancement grading was established as follows: Grade 0 indicated no enhancement or similar to the adjacent normal intracranial artery wall; Grade 2 was comparable to or exceeded the physiological enhancement of the pituitary gland; Grade 1 denoted an enhancement level intermediate to the aforementioned grades.

## Statistical analysis

Statistical analyses were conducted using IBM SPSS software (version 22.0, IBM, Armonk, NY), with variables presented as mean ± standard deviation or as a percentage (%). The intraclass correlation coefficient (ICC) assessed the reliability of plaque feature measurements. The Kolmogorov-Smirnov and Levene tests were initially applied to assess the normality and homogeneity of variance of the parameters, respectively. The T-test compared differences between two groups for normally distributed variables, while the Mann-Whitney U test was utilized for skewed data, and the chi-square test for categorical variables. Significant clinical or plaque feature parameters were analyzed using the variance inflation factor (VIF) to detect multicollinearity. Binomial logistic regression models identified independent risk factors for positive remodeling (PR), with odds ratios (ORs) and 95% confidence interval (CI) calculated. Diagnostic performance was described using receiver operating characteristic (ROC) curves and area under curve (AUC) values.

## Results

A total of 84 patients (mean age 58.07±1.35 years, 78.6% male) were categorized into PR (n=28, 33.3%) or NPR (n=56, 66.4%) group based on the remodeling ratio (RR). Within the NPR group, 54 exhibited negative remodeling and 2 showed no remodeling. Demographic data are detailed in the accompanying Table. 2. No significant differences were observed in sex, age, or scan timing post-symptom onset between groups.

**Table 2.**
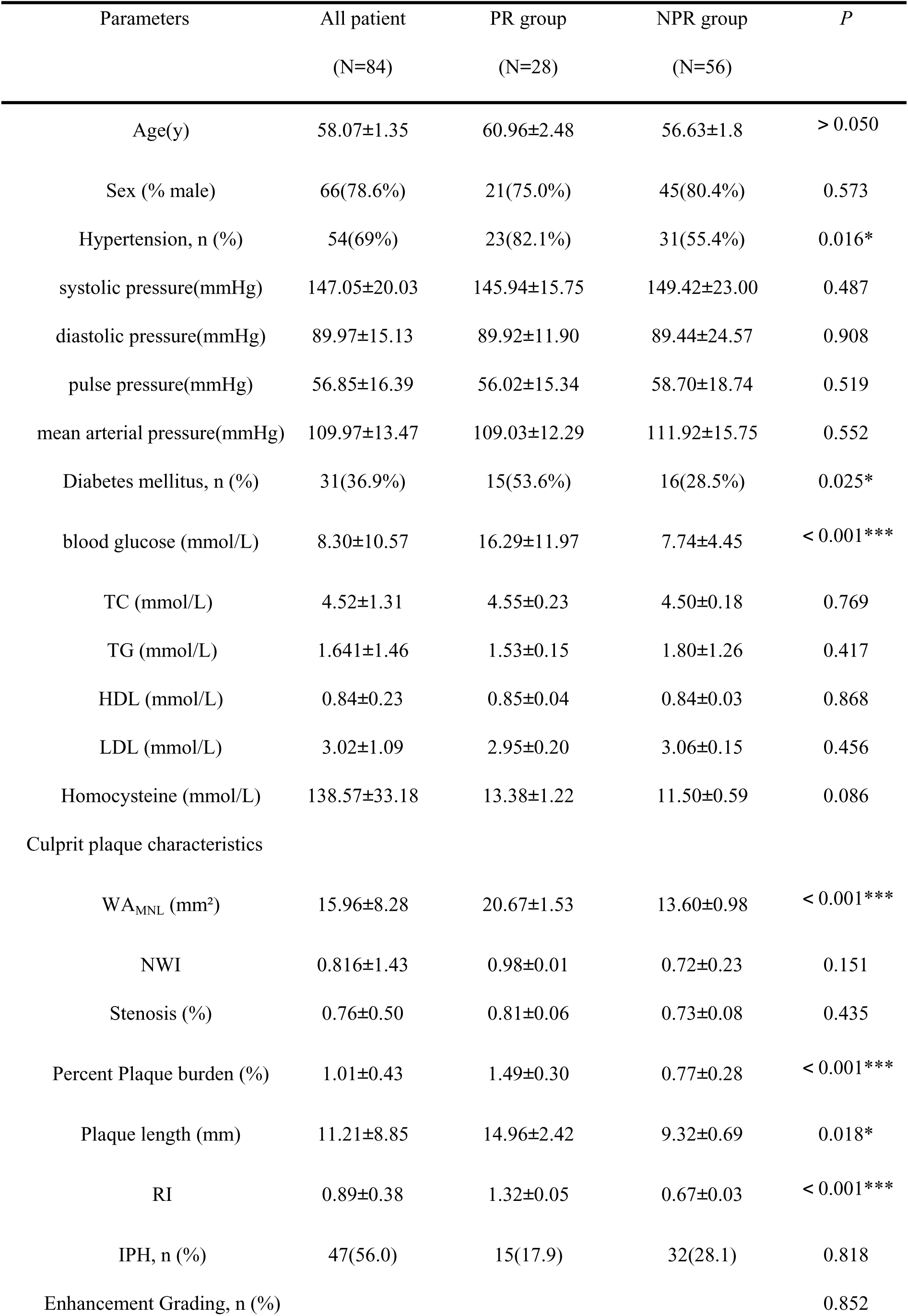
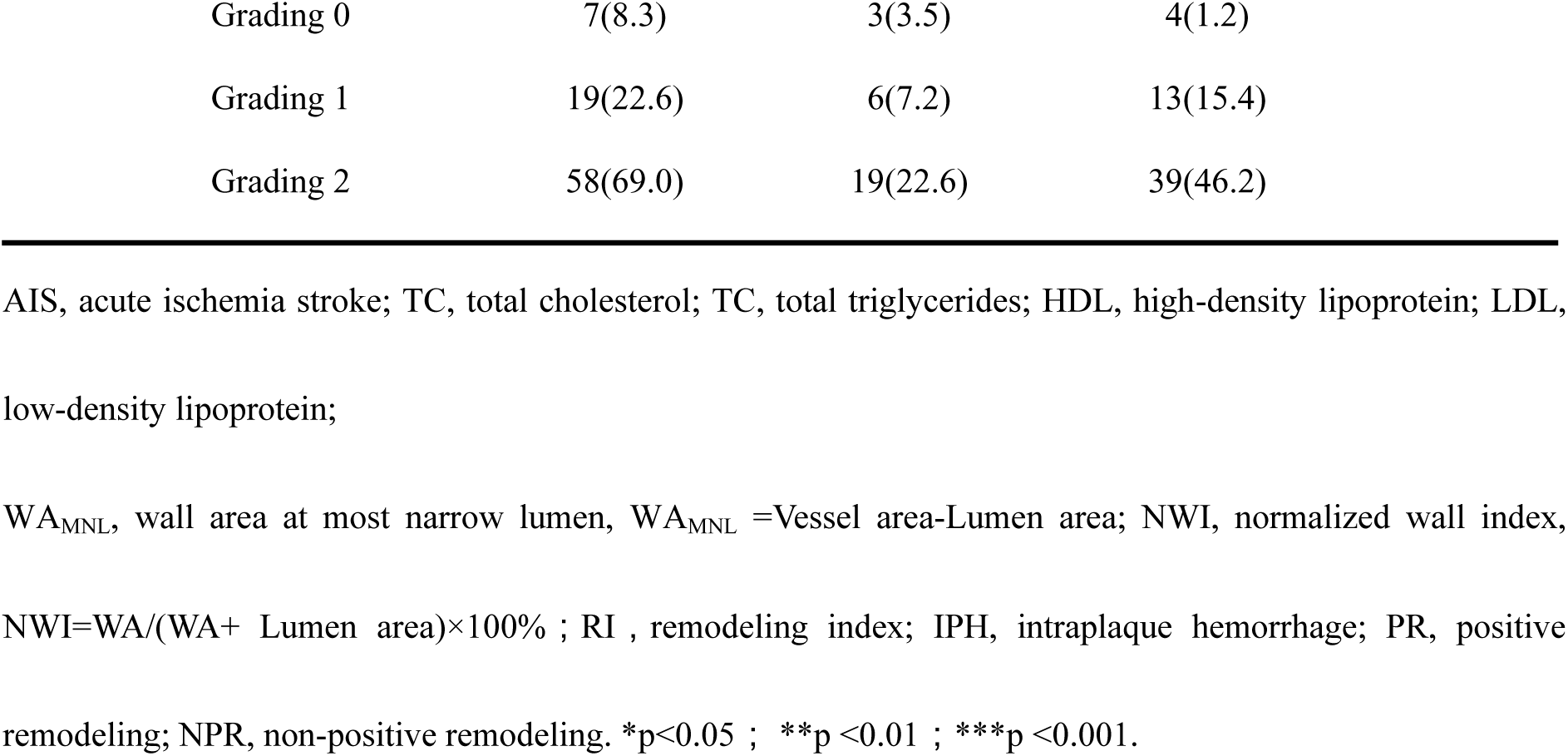
Characteristics of AIS patients with different patterns of vessel remodeling.

| Parameters | All patient<br>(N=84) | PR group<br>(N=28) | NPR group<br>(N=56) | *P* |
| --- | --- | --- | --- | --- |
| Age(y) | 58.07±1.35 | 60.96±2.48 | 56.63±1.8 | > 0.050 |
| Sex (% male) | 66(78.6%) | 21(75.0%) | 45(80.4%) | 0.573 |
| Hypertension, n (%) | 54(69%) | 23(82.1%) | 31(55.4%) | 0.016* |
| systolic pressure(mmHg) | 147.05±20.03 | 145.94±15.75 | 149.42±23.00 | 0.487 |
| diastolic pressure(mmHg) | 89.97±15.13 | 89.92±11.90 | 89.44±24.57 | 0.908 |
| pulse pressure(mmHg) | 56.85±16.39 | 56.02±15.34 | 58.70±18.74 | 0.519 |
| mean arterial pressure(mmHg) | 109.97±13.47 | 109.03±12.29 | 111.92±15.75 | 0.552 |
| Diabetes mellitus, n (%) | 31(36.9%) | 15(53.6%) | 16(28.5%) | 0.025* |
| blood glucose (mmol/L) | 8.30±10.57 | 16.29±11.97 | 7.74±4.45 | < 0.001*** |
| TC (mmol/L) | 4.52±1.31 | 4.55±0.23 | 4.50±0.18 | 0.769 |
| TG (mmol/L) | 1.641±1.46 | 1.53±0.15 | 1.80±1.26 | 0.417 |
| HDL (mmol/L) | 0.84±0.23 | 0.85±0.04 | 0.84±0.03 | 0.868 |
| LDL (mmol/L) | 3.02±1.09 | 2.95±0.20 | 3.06±0.15 | 0.456 |
| Homocysteine (mmol/L) | 138.57±33.18 | 13.38±1.22 | 11.50±0.59 | 0.086 |
| Culprit plaque characteristics |  |  |  |  |
| WA<sub>MNL</sub> (mm<sup>2</sup>) | 15.96±8.28 | 20.67±1.53 | 13.60±0.98 | < 0.001*** |
| NWI | 0.816±1.43 | 0.98±0.01 | 0.72±0.23 | 0.151 |
| Stenosis (%) | 0.76±0.50 | 0.81±0.06 | 0.73±0.08 | 0.435 |
| Percent Plaque burden (%) | 1.01±0.43 | 1.49±0.30 | 0.77±0.28 | < 0.001*** |
| Plaque length (mm) | 11.21±8.85 | 14.96±2.42 | 9.32±0.69 | 0.018* |
| RI | 0.89±0.38 | 1.32±0.05 | 0.67±0.03 | < 0.001*** |
| IPH, n (%) | 47(56.0) | 15(17.9) | 32(28.1) | 0.818 |
| Enhancement Grading, n (%) |  |  |  | 0.852 |

|  |  |  |  |
| --- | --- | --- | --- |
| Grading 0 | 7(8.3) | 3(3.5) | 4(1.2) |
| Grading 1 | 19(22.6) | 6(7.2) | 13(15.4) |
| Grading 2 | 58(69.0) | 19(22.6) | 39(46.2) |

AIS, acute ischemia stroke; TC, total cholesterol; TG, total triglycerides; HDL, high-density lipoprotein; LDL, low-density lipoprotein;
WA<sub>MNL</sub>, wall area at most narrow lumen, WA<sub>MNL</sub> =Vessel area-Lumen area; NWI, normalized wall index, NWI=WA/(WA+ Lumen area)×100% ; RI , remodeling index; IPH, intraplaque hemorrhage; PR, positive remodeling; NPR, non-positive remodeling. \*p<0.05 ; \*\*p <0.01 ; \*\*\*p <0.001.

## Differentiation between PR and NPR groups

The distribution of remodeling patterns among culprit plaques in AIS patients showed no significant difference(p=0.091), with 47 non-PR plaques in the MCA and 9 plaques in the BA, compared to 19 PR plaques in the MCA and 9 in the BA, as showed in Fig.3.

**Fig.1.**
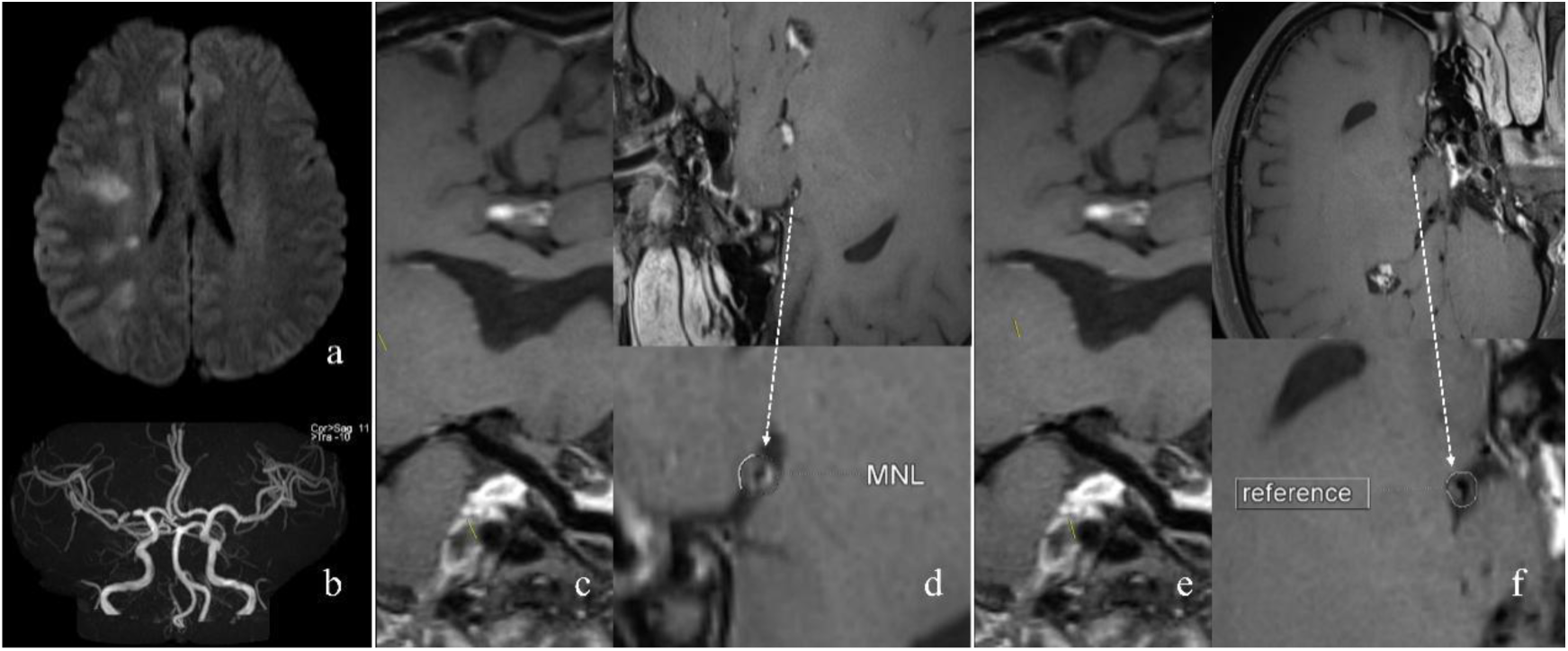
Positive remodeling pattern in patients with AIS. (a)DWI. (b) TOF MRA MIP. (c) and (e) were MNL site and reference site on curved-planar reconstructed post enhancement T1WI. (d) MNL site on the cross section of vessel (arrow) and measurement of vessel area. (f) Reference site on the cross section of vessel (arrow) and measurement of vessel area. AIS, acute ischemic stroke; DWI, diffuse weighted imaging; TOF, time of fly; MRA, magnetic resonance angiography; MIP, maximum intensity projection; MNL, most narrow lumen.

**Fig.2.**
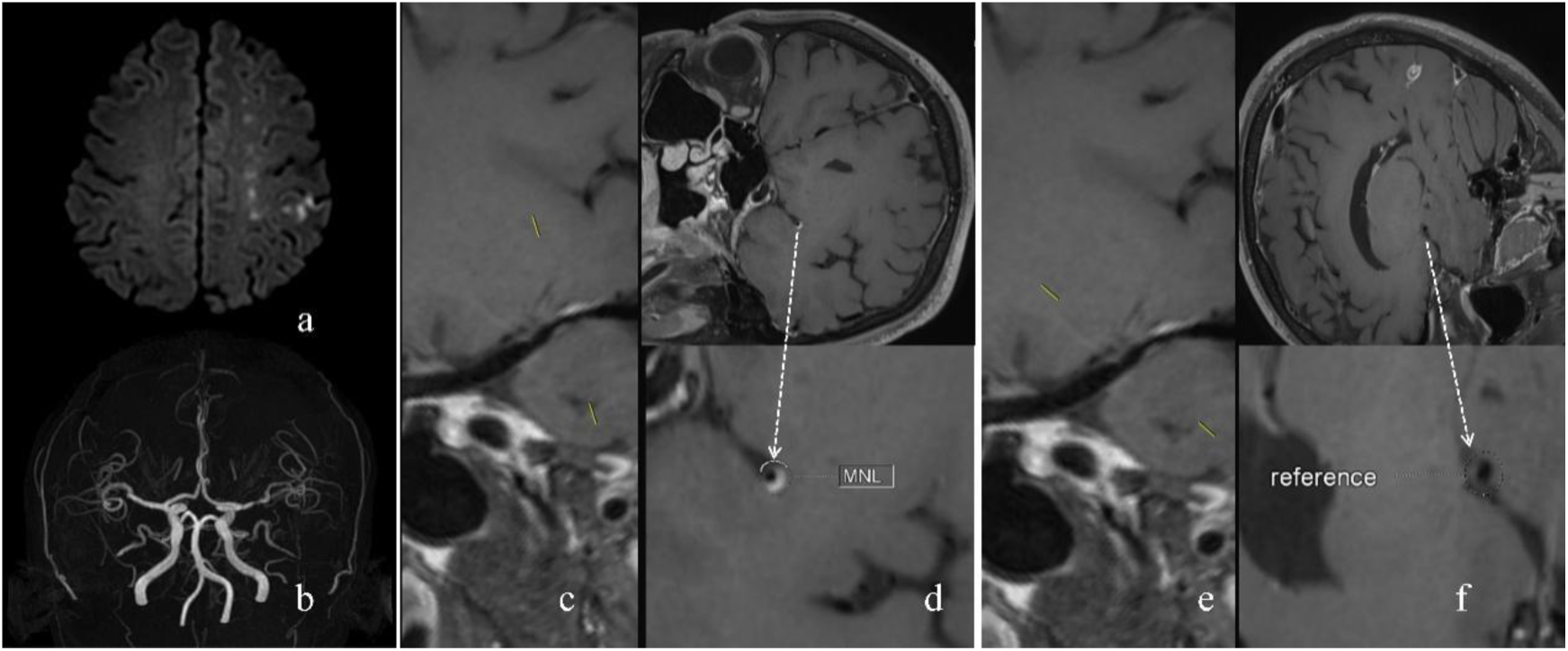
None positive remodeling pattern in patients with AIS. (a)DWI. (b) TOF MRA MIP. (c) and (e) were MNL site and reference site on curved-planar reconstructed post enhancement T1WI. (d) MNL site on the cross section of vessel (arrow) and measurement of vessel area. (f) Reference site on the cross section of vessel (arrow) and measurement of vessel area. AIS, acute ischemic stroke; DWI, diffuse weighted imaging; TOF, time of fly; MRA, magnetic resonance angiography; MIP, maximum intensity projection; MNL, most narrow lumen.

**Fig.3.**
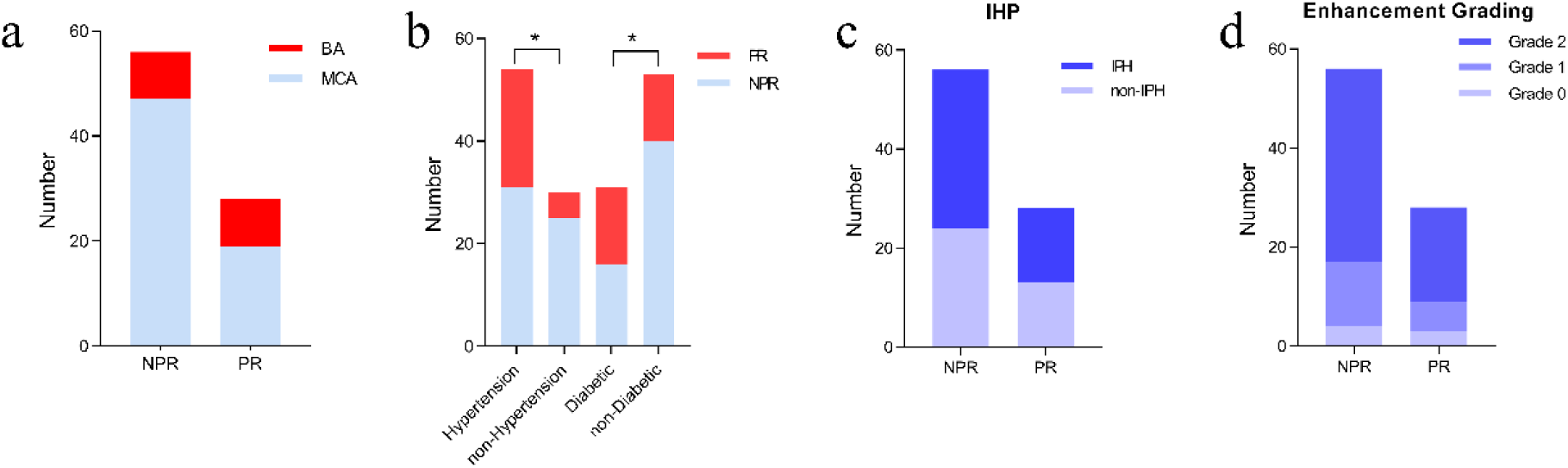
culprit plaque location (b), Clinical factors (a), and plaque features (c, d) between PR and NPR in the patients with AIS. BA, basilar artery; MCA, cerebral middle artery; IPH, intraplaque hemorrhage; PR, positive remodeling NPR, none positive remodeling; AIS, acute ischemic stroke.

Notable differences between PR and NPR groups included percent plaque burden (1.49%±0.30% vs 0.77%±0.28%, P<0.001), plaque length (14.96±2.42mm vs 9.32±0.69mm, P=0.018), and the prevalence of high blood pressure (HBP) (82.1% vs 55.4%, p=0.016) and diabetes mellitus (53.57% vs 28.57%, p=0.025). However, no significant differences were found between the two groups regarding intraplaque hemorrhage (IPH), enhancement score, systolic and diastolic pressures, pulse pressure, and mean arterial pressure, as showed in Fig.3 and Fig.4.

**Fig.4.**
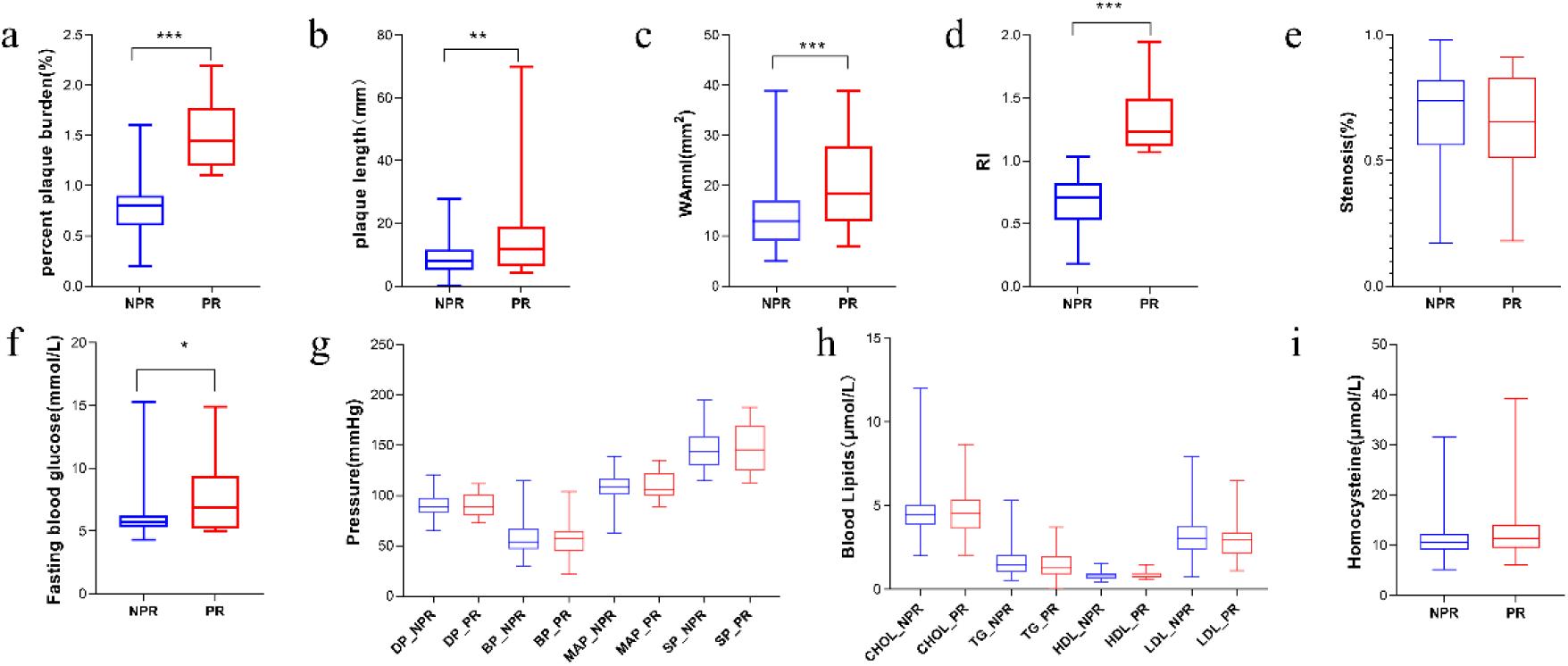
Box plots showing comparison of culprit plaque features(a-e) and clinical factors (f-i) between PR and NPR in the patients with AIS. PR, positive remodeling NPR, none positive remodeling; AIS, acute ischemic stroke.

## Correlations between parameters and RI

Percent plaque burden showed strong correlations with RI (r=0.95, 0.98, P<0.001), while WA_MNL_, FBG were moderately and weekly related to RI (r=0.57, 0.27, P<0.001, 0.035), as shown in Fig.5.

**Fig 5.**
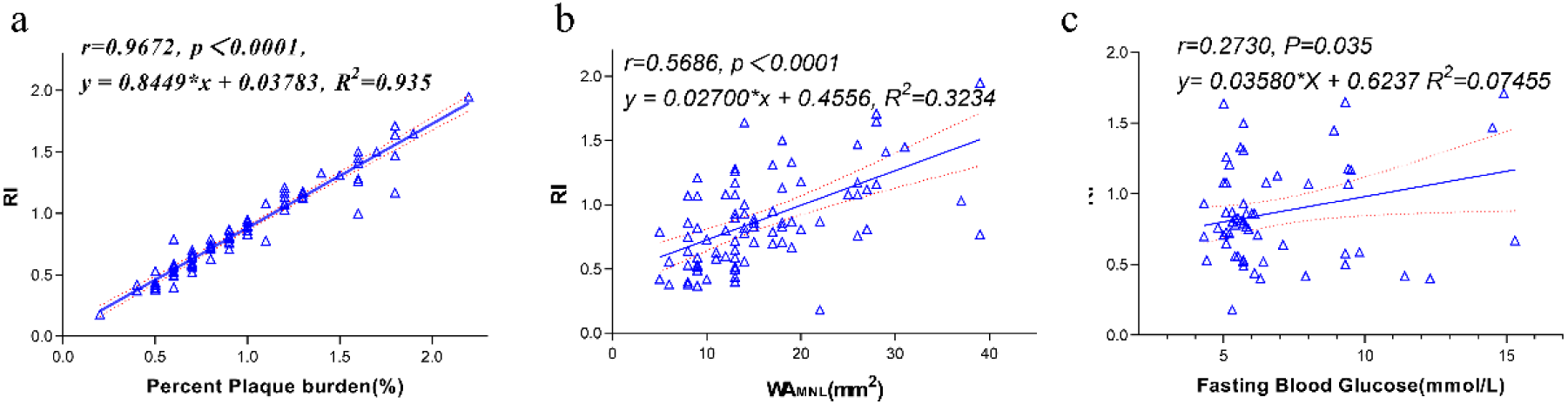
Correlation of plaque measurements (a, b) and fasting blood glucose(c) with RI. WAMNL, wall area at the most narrowed site; RI, remodeling index.

## Independent factors of positive remodeling

Adjusting for clinical factors, binary logistic regression analysis revealed percent plaque burden as an only independent predictor of PR (OR 4.19, per 10% increase; 95% CI, 1.79-9.81, p=0.001), the AUC of the percent plaque burden was 0.984, the sensitivity was 100%, and the specificity was 94.64%, as shown in Table. 3 and Fig.6.

**Table 3.** Association between investigated parameters with positive remodeling in patients with AIS comparing to non-positive remodeling.

| Variable | OR ( 95%CI ) | Unadjusted P | Adjusted OR ( 95%CI ) | Adjusted P* |
| --- | --- | --- | --- | --- |
| Plaque burden (per 10%) | 3.82(1.87-7.79) | < 0.001 | 4.19(1.79-9.81) | 0.001 |
| Plaque length (mm) | 1.10(1.02-1.19) | 0.015 | 1.00(0.84-1.19) | 0.968 |
| Hypertension | 3.7(1.23-11.16) | 0.020 | 0.054 ( 0.00-1.44 ) | 0.082 |
| Diabetes | 2.89(1.12-7.40) | 0.028 | 0.511 ( 0.04-7.16 ) | 0.618 |

Adjusted P\*, outcomes adjusted by other variables. AIS acute ischemia stroke; OR, odds ratio; CI, Confidence interval.

**Fig. 6.**
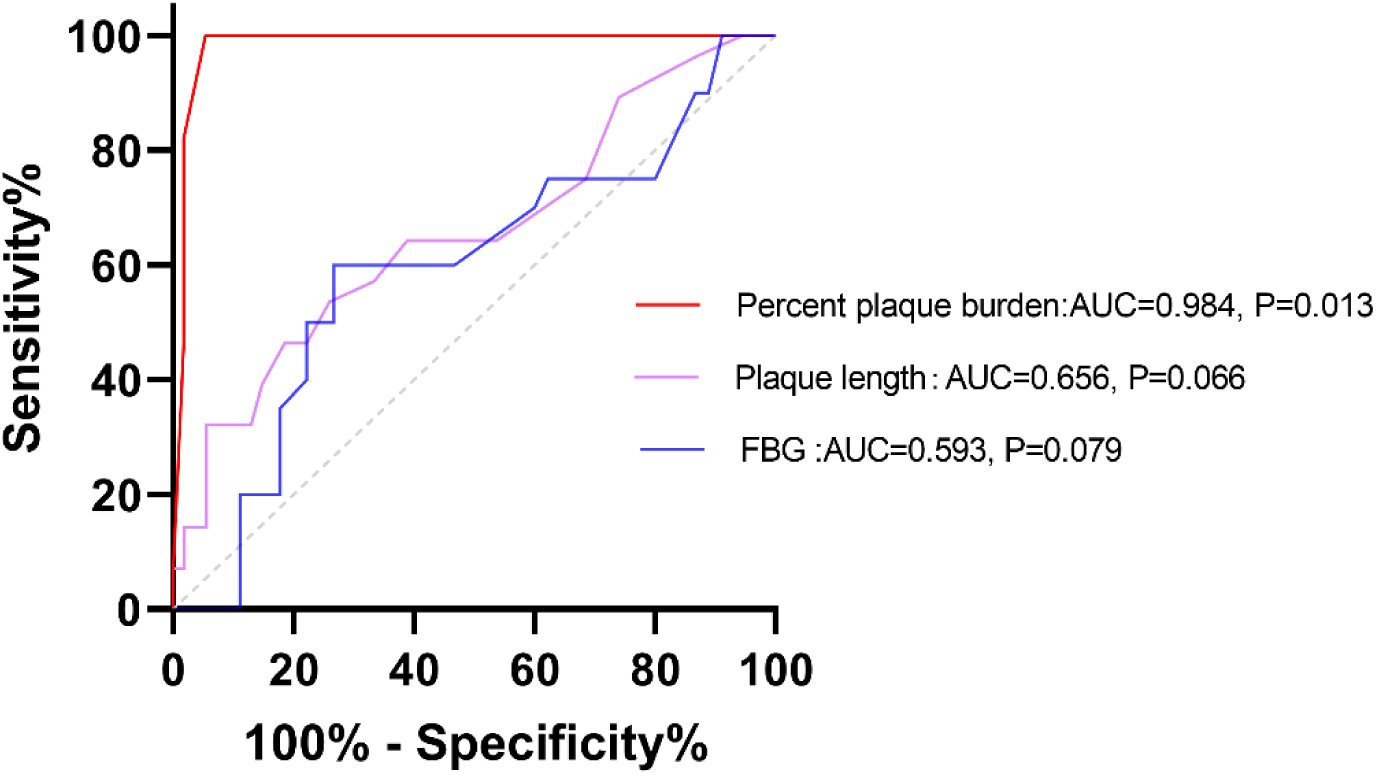
Receiver operating characteristic curves combining three parameters were compared to distinguish between PR and NPR in AIS. FBG, fasting blood glucose; PR, positive remodeling NPR, none positive remodeling; AIS, acute ischemic stroke; AUC, area under curve.

## Discussion

In this prospective observational study, the model was initially utilized to explore factors associated with PR of ICAS culprit plaques in AIS, using in vivo high-resolution vessel wall imaging (HR-VWI). The study compared the clinical status, qualitative imaging features, and quantitative imaging parameters between PR and non-PR groups, which included 28 with positive remodeling, 54 with negative remodeling, and 2 with no remodeling. The predominant remodeling pattern for both the basilar artery (BA) and middle cerebral artery (MCA) was negative, yet there was a nearly twofold higher prevalence of PR. Significant differences were observed in percent plaque burden, plaque length, and history of high blood pressure (HBP) and diabetes mellitus between the two groups. After adjusting for multiple factors, percent plaque burden emerged as the sole risk factor related to PR, with an AUC of 0.984, effectively distinguishing PR from non-PR. These findings indicate that local percent plaque burden may be crucial for PR and suggest that noninvasive assessment of plaque morphology could serve as a useful marker for stratifying atherosclerotic risk.

Several lines of evidence from prior observational studies on symptomatic plaques^28,29^ have suggested that outward remodeling may be associated with plaque instability, potentially leading to stroke. Interestingly, in our study, the predominant remodeling pattern of culprit plaques in the BA and MCA is negative remodeling. This discrepancy may be attributable to variations in cohort selection and research objectives. Our study focused on culprit plaques implicated in AIS, confirmed by high signal intensity on DWI, which were not specified in earlier studies, and we now identified imaging features and clinical factors influencing remodeling. Whereas most former studies distinguished symptomatic (presumed ischemic stroke or TIA) from asymptomatic arteries, or stenotic from non-stenotic arteries, to assess plaque stability.

Employing non-invasive in vivo MRI, we determined that plaques undergoing PR exhibited a significantly larger and longer burden than those with NPR. The percentage plaque burden displayed strong correlations with RI, and emerged as an independent predictive factor strongly associated with PR of culprit lesions. This was in line with earlier findings from aortic plaque studies in rabbit models^14^, which indicated a strong relationship between plaque burden, positive remodeling, and plaque disruption, characterized by histological features such as a thin fibrous cap and a dense lipid core.

Contrary to expectations, intra-plaque hemorrhage (IPH) and enhancements in AIS patients, indicative of vulnerable plaque disruption followed by cute thrombus formation leading to rapid plaque expansion, were not associated with positive remodeling. Kristinel^21^ also observed that RR did not differ between plaques with IPH and those without IPH in patients with recent ischemic events. A plausible explanation is that the remodeling responses to plaque burden represents an ongoing process in reaction to persistent atherosclerotic changes and is not influenced by short-term morphological alterations of the plaque.

The terminology used to describe plaque burden is contentious and lacks uniformity. Changes in the wall area were described as “plaque size” or “plaque area” in prior researches^17,28,29^ and calculated as WA_MNL_ −WA_reference_. Yet, changes in wall area encompass not only increases in the plaque mass but also extensive alterations in arterial circumference, affecting each arterial layer, known as vascular remodeling. In line with the precise classification by Mulvany et al.^6^, changes in vessels are categorized based on lumen diameter alterations (inward or outward) and wall area changes (increased=hypertrophic, decreased = hypotrophic, no change=eutrophic). The arterial changes in our positive remodeling (PR) group are characterized by a decrease in lumen area but an increase in wall area (hypertrophic). Conversely, most vessels in the non-PR group displayed hypotrophic inward remodeling, marked by reductions in both lumen and vessel size, which potentially due to constraints imposed by a stiff outer layer^30^. Precent plaque burden, defined as the ratio of the lesion site wall area to the reference site wall area, more accurately reflects vascular deformation resulting from long-term plaque evolution and is closely associated with the RI (p < 0.001), consistent with previous studies^28,31^.

Additionally, our findings indicate that a history of hypertension and diabetes may enhance and accelerate PR. Both conditions have been implicated in ICAS and are known to accelerate atherosclerotic plaque formation. Research on arterial remodeling due to hypertension and diabetes has predominantly focused on the coronary and carotid arteries, yet there is no consensus on their impact on arterial remodeling in various vascular territories. A combined ultrasound and HR-MRI study^19^ of carotid arteries revealed that complex plaques (American Heart Association [AHA] stages IV-VIII) were more prevalent in patients with hypertension, dyslipidemia, and diabetes and were associated with positive remodeling compared to less complex plaques (AHA stages I-III). Conversely, a 9-month follow-up study of nontreated mild lesions using intracoronary ultrasound imaging^32^ in patients with type 2 diabetes showed a bidirectional pattern of vascular remodeling in atherosclerotic plaques, with one-third demonstrating shrinkage and two-thirds non-shrinkage. However, Beaussier et al.^33^ and Laugesen et al.^34^ reported that carotid artery walls in hypertensive and diabetic patients are more likely to undergo negative remodeling, characterized by a decrease in vascular area as plaques develop, and poor glycemic control, as measured by glycated albumin, was linked to negative coronary artery remodeling^35^. In our study, over 80% of patients in the PR group had a history of hypertension, yet no significant differences were observed in pulse pressure, mean arterial pressure, systolic pressure, and diastolic pressure between the two groups, which could be attributed to cerebrovascular autoregulation or the use of antihypertensive therapies. Furthermore, VSMC proliferation is considered a key factor in arterial wall thickening. Compared to extracranial arteries, the unique structure of intracranial arteries, featuring one or two layers of VSMCs, may result in relatively insufficient VSMC proliferation. A high prevalence of diabetes mellitus history was associated with cerebrovascular positive remodeling, and the FBG levels in the PR group were higher than those in the non-PR group, but FBG was only weakly correlated with the RI (r = 0.27, P = 0.035).

## Limitation

First, our study did not account for the impact of vessel tapering on measurement, designating the reference site as the segment with the least atherosclerotic disease and overlooking potential remodeling in these segments. This oversight may lead to underestimation or overestimation of our findings. Second, we only included initial clinical data on blood pressure and biomarkers collected after patients were admitted to the emergency department, without subsequent follow-up data post-symptom remission, which introduces potential selection bias. The variance in the duration of hypertension, treatment duration, and patient compliance, which could influence outcomes, was not ascertained. We also did not gather comprehensive parameters related to glycemic control, such as glycated albumin, glycated hemoglobin, and advanced glycation end-products, which may provide a more accurate assessment than FBG alone. Lastly, arterial remodeling is an ongoing biological process within ICAS plaques. Ruptured plaques with HIP and enhancement, followed by late healing, could influence the remodeling trajectory, therefore, longitudinal studies are necessary to monitor this process and ascertain its impact on long-term patient outcomes.

## Conclusion

HR-VWI enables non-invasive detection of remodeling in ICAS, potentially leading to novel methods for evaluating the influence of plaque morphological characteristics and clinical parameters on cerebrovascular remodeling patterns, thereby improving cardiovascular risk stratification. A high burden of culprit plaque is an independent predictor of positive remodeling.

## Data Availability

The authors confirm that the data supporting the findings of this study are available within the article and supplementary materials.For further data requests, please contact the corresponding authors.

## Non-standard Abbreviations and Acronyms

AIS: acute ischemic stroke
AUC: area under curve
BA: basilar artery
DWI: diffuse weighted imaging
FBG: fasting blood glucose
ICAS: intracranial atherosclerosis
IPH: intraplaque hemorrhage
MRA: magnetic resonance angiography
MCA: middle cerebral artery
MIP: maximum intensity projection
MNL: most narrow lumen
NPR: none positive remodeling
PR: positive remodeling
RI: remodeling index
TOF: time of fly
VSMC: vascular smooth muscle cell

## Acknowledgments

We are grateful to Doctor Li-ping Ma for her help with conception and design, Doctor Di-Hao Xu for his advice on ROI measurements, and Doctor Jing-wen Xie for the help with imaging scan.

## Disclosures

The authors declare that they have no conflicts of interest.

This manuscript has been submitted as conference abstract in RSNA-2023(Scientific Poster Sessions presentation: W5A-SPNR-4).

## Supplemental Material

**Fig.S1.**
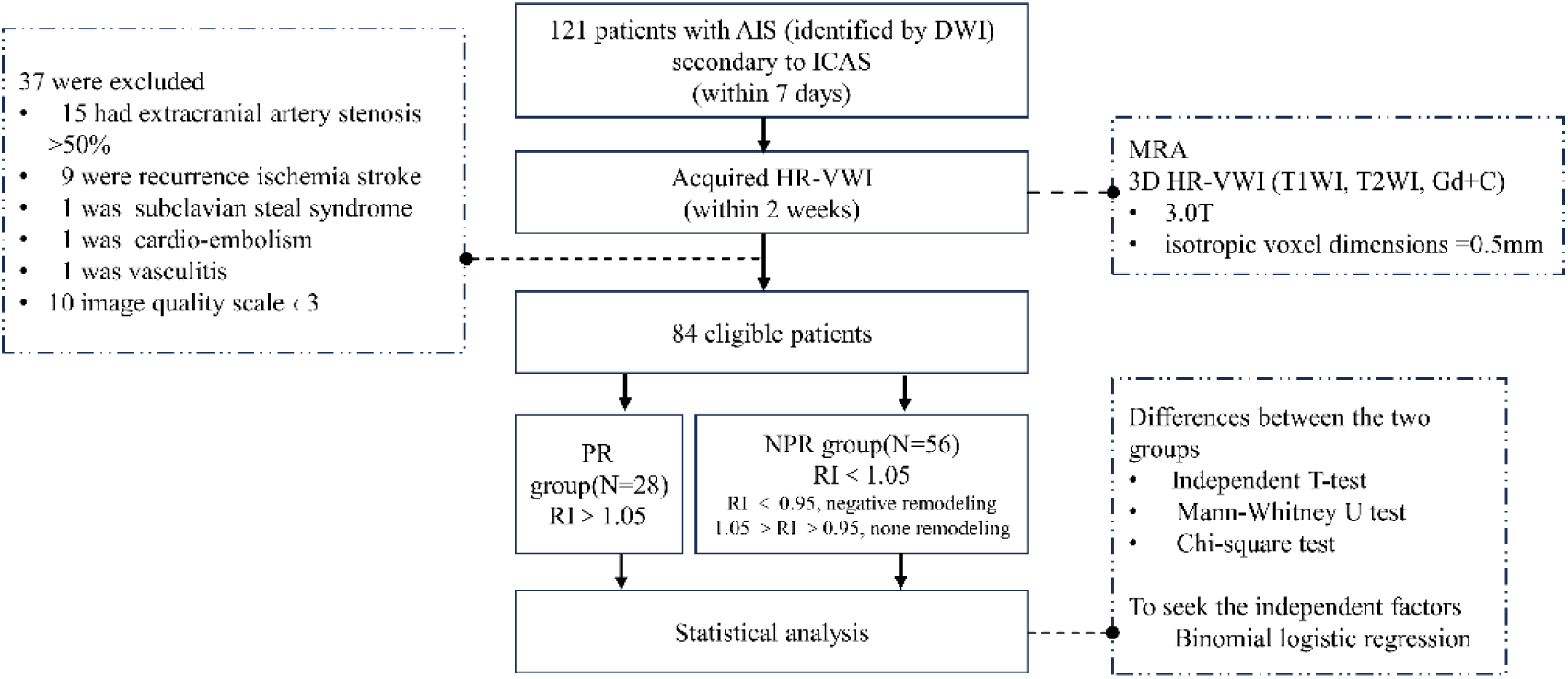
Flow chart of the study. AIS, acute ischemic stroke; ICAS, intracranial atherosclerosis; HR-VWI, High-resolution vascular wall imaging; DWI, diffuse weighted imaging; RI, remodeling index, calculated as vessel area MNL /vessel area reference; PR, positive remodeling NPR, none positive remodeling.

